# Impact of Cardiomyopathy and Arrhythmia Genetic Testing on Clinical Management Decisions

**DOI:** 10.64898/2026.07.22.26358740

**Authors:** Ana Morales, Yi-Lee Ting, Brianna Bucknor, C. Anwar A. Chahal, Emily Higgs, Daniel P. Judge, Anjali T. Owens, Jessica Wang, Mohamad Alkhayat, Naomi Barker, Megan Betts, Jessica L. Chowns, Rebecca M. Eberly, Edward D. Esplin, Lily Hoffman-Andrews, Ajit Koduri, Ajith Nair, Arun Padmanabhan, Vasanth Vedantham, Julianne Wojciak, Elizabeth M. McNally

**Affiliations:** Department of Genomic Health, Geisinger, Danville, PA, USA; Translational Health Sciences Program, School of Medicine and Health Sciences, The George Washington University, Washington, DC, USA; Former affiliation: Invitae Corporation (now part of Labcorp), San Francisco, CA; Labcorp (formerly Invitae Corporation), San Francisco, CA, USA; Wellspan Health, Center for Inherited Cardiovascular Diseases, Precision Medicine, York, PA, USA; Department of Cardiovascular Medicine, Mayo Clinic, Rochester, MN, USA; William Harvey Research Institute, Queen Mary University of London, London, UK; Cardiovascular Genetics Center, Cardiology Division, University of California, San Francisco, CA, USA; Medical University of South Carolina, Charleston, SC, USA; University of Pennsylvania Perelman School of Medicine, Philadelphia, PA, USA; University of California, Los Angeles, CA, USA; Penn Medicine Division of Cardiovascular Medicine, Perelman School of Medicine at the University of Pennsylvania, Philadelphia, PA, USA; Baylor College of Medicine, The Texas Heart Institute, Houston, TX, USA; Cardiovascular Research Institute, University of California, San Francisco, CA, USA; Center for Genetic Medicine, Northwestern University Feinberg School of Medicine, Chicago, IL, USA

**Keywords:** Cardiomyopathy, genetic testing, genetics-informed care

## Abstract

**Background:** Genetic testing for cardiomyopathy and arrhythmia (CM/ARRH) provides diagnostic information and informs screening for at-risk relatives. Clinical guidelines recommend genetic testing for these conditions; however, data on how genetic test results influence clinical management recommendations are limited. Here, we determined the frequency of cardiologist-recommended management changes for patients following CM/ARRH genetic testing.

**Methods:** This was a retrospective cross-sectional study of patients referred for multigene panel testing between April 2016 and April 2024. Genetically-experienced cardiologists at multicenter academic clinical practices were recruited for participation to complete surveys indicating clinical decision making on patients who had genetic testing. Cases for review were randomly selected to have both positive and non-positive results.

**Results:** Among 249 patients (138 positive, 111 non-positive), 136 (54.6%) received clinical management recommendations for their own or their at-risk relatives’ care. Of these, 75 (55.1%) received recommendations for the patient’s own care, most frequently additional diagnostic tests/procedures (n=33). Compared to non-positive results, patients with positive results were more likely to receive recommendations for their own management (66/138, 47.8% vs 9/111, 8.1%; P<0.00001). Patients with positive results in arrhythmogenic cardiomyopathy genes had 263% higher odds of recommended management changes compared to those with *TTN* (OR=3.63, CI:1.40-9.84, P=0.009).

**Conclusions:** The results from CM/ARRH genetic testing on affected patients influenced cardiologists’ medical decision making and management recommendations. Additional research is needed to evaluate how genetic testing for CM/ARRH impact health outcomes.

## INTRODUCTION

Genetic cardiomyopathy (CM) and arrhythmia (ARRH) are prevalent, clinically heterogeneous disorders that have an increased risk for sudden death and heart failure. Genetic testing for these conditions is widely available through both academic and commercial laboratories. Broad panel testing encompassing both CM and ARRH genes reveals clinically relevant variants in 1 in 5 patients.^1,2^

Clinical guidelines recommend genetic testing for cardiomyopathy since the results help guide diagnosis, inform clinical management changes like arrhythmia risk monitoring, and refine surveillance needs for at-risk family members.^3–7^ Despite professional society recommendations, it is estimated that only 1% of patients with CM or ARRH receive genetic testing, indicating barriers to genetic testing uptake.^8,9^ Barriers to cardiac genetic testing utilization include clinician, patient, and institutional factors.^10^ In one study, 45% of non-genetic cardiologists did not feel confident in making clinical recommendations based on genetic test results.^11^ In another study, 43% of cardiologists expressed insufficient confidence in interpreting results; however, cardiologists who more frequently order genetic testing are less likely to express this concern.^12^

Studies reporting implications of genetic diagnosis for CM/ARRH have focused mainly on diagnostic yield and family member cascade testing,^1,13–20^ with less attention paid to genetic-informed management for affected patients. Linking specific medical actions to receipt of genetic testing results is complicated by the temporally long clinical course of cardiomyopathy, where many management adjustments occur independent of genetic testing results. A first step to assess the potential clinical utility of an intervention is to query its role in clinical decision making. This study addressed this knowledge gap by surveying cardiologists with expertise in cardiac genetic testing about their clinical recommendations following the return of genetic testing results. We reasoned that cardiologists familiar with genetic testing for CM/ARRH were less likely to be inhibited in ordering and interpreting genetic testing results and represented an optimized setting for this analysis.

## METHODS

### Ethics approval

The study was approved by the Western Institutional Review Board (tracking number 20210520). No personal health information was collected on the survey; therefore, the study was considered exempt. Data were excluded for patients who opted out of data use for research purposes or requested data deletion after completion of genetic testing.

### Data availability statement

The data that supports the findings of this article include proprietary and protected information, which cannot be made publicly available in its entirety. Selected deidentified data may be made available upon reasonable request from the authors. YT and BB had full access to the data in the study and took responsibility for its integrity and data analysis.

### Study design

A retrospective survey was conducted to investigate how genetically-experienced cardiologists use genetic testing to inform disease management of patients affected with CM/ARRH. To determine cohort size for 80% power to detect effects of a medium size, we assumed approximately 50% of patients with a positive result had a management change recommendation.

### Inclusion criteria

#### Cardiology providers

Research personnel from a commercial laboratory reached out to cardiology providers who had regularly ordered the multi-gene next-generation sequencing CM/ARRH panel (Supplemental Table S1) and invited them to participate in the study. These clinicians were selected because they were deemed “experienced” based on ordering volume and authorship in a professional society guideline, peer-reviewed diagnostic cardiovascular genetics-focused publication, or who worked with an expert at the same location. A minimum of 50 and 150 test orders was required from guideline and other literature authors, respectively. This approach identified 15 cardiologists, who were invited to the study. Of these, 11, from 7 institutions, agreed to participate (Figure 1). Cardiologists were not compensated for participation in this research.

**Figure 1.**
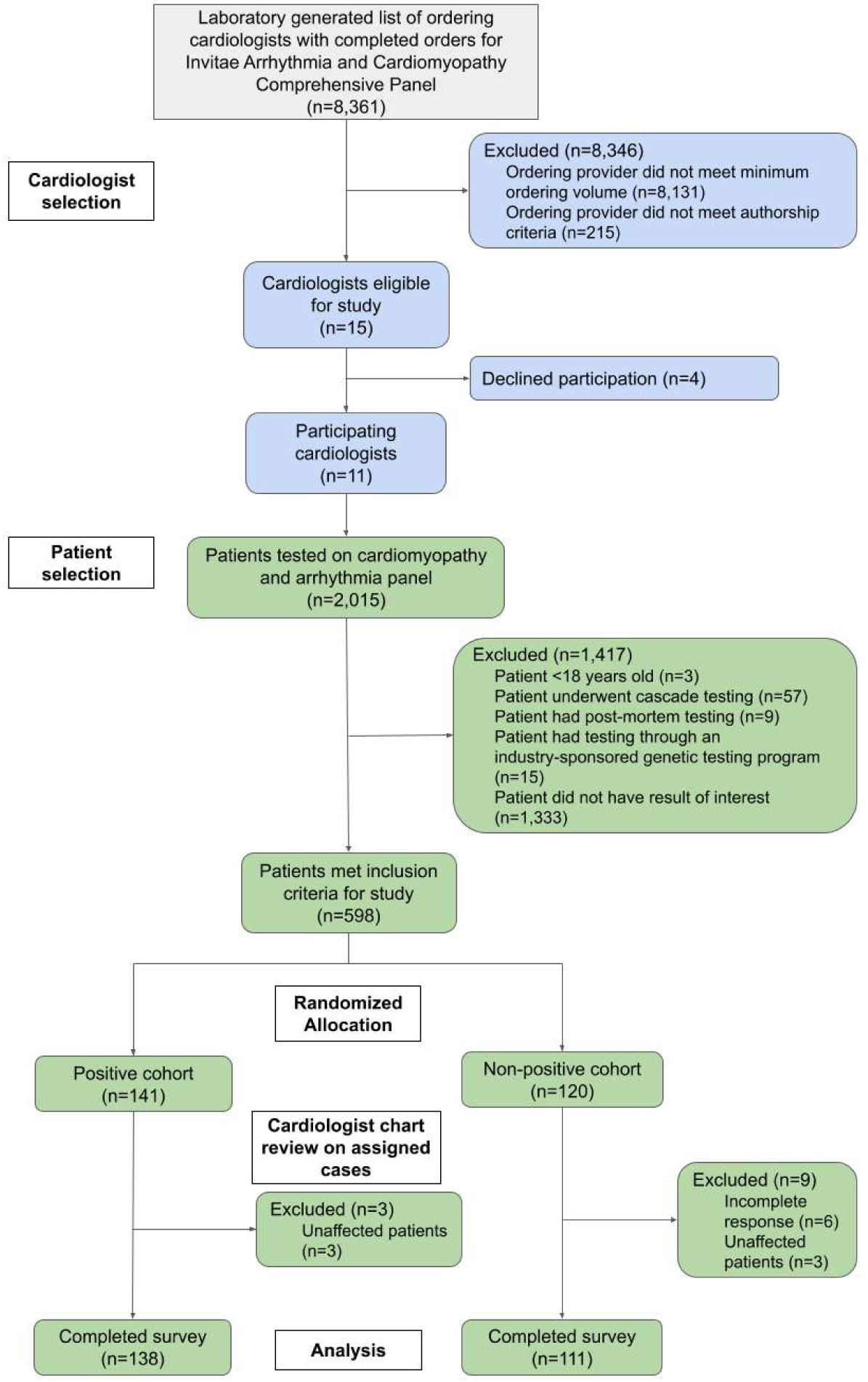
Flowchart for participating cardiologists and cohort development. Cardiologists were invited to participate based on their clinical expertise in genetic testing for cardiomyopathy and arrhythmia conditions. After agreeing to participate, cardiologists completed case report forms from a retrospective chart review on randomly selected patients who received positive results in an arrhythmogenic cardiomyopathy gene plus *TTN* or non-positive results.

#### Cases with CM/ARRH

For case reviews, a retrospective search was made for patients who were ≥18-years-old at the time of testing and had the CM/ARRH panel ordered between April 2016 and April 2024. The following groups were excluded from analysis: patients who only underwent cascade testing, post-mortem status documented in the test requisition form, and industry-sponsored genetic testing programs. Additionally, to eliminate confounding from the possibility of management change recommendations made for patients with VUS that a clinician may have decided to manage as a positive result,^21^ those with only a VUS in an established CM/ARRH autosomal dominant gene were excluded.

For each participating institution, a random selection of patients with positive results in a specific gene of interest and non-positive results was made. The positive group included individuals who had a pathogenic/likely pathogenic (P/LP) variant in one of the arrhythmogenic cardiomyopathy (ACM) genes referenced in the 2019 Heart Rhythm Society (HRS) expert consensus statement on A CM:^22^ *BAG3*, *DES*, *DSC2*, *DSG2*, *DSP*, *FLNC*, *JUP*, *LDB3*, *LMNA*, *NKX2-5*, *PKP2*, *PLN*, *RBM20*, *SCN5A*, *TMEM43*These genes, associated with dilated and arrhythmogenic cardiomyopathy phenotypes, were selected because arrhythmia risk monitoring and specialized management is recommended for patients with a molecular diagnosis in these genes. *TTN* was also included as a gene of interest based on participating cardiologists’ expert input and published evidence documenting an arrhythmogenic phenotype associated with this gene.^23,24^ The non-positive group included the following patients with non-actionable results: those with a negative result (no reportable variants), only a carrier result (heterozygous P/LP variant in an autosomal recessive gene), or a variant of uncertain significance (VUS) in a gene of uncertain significance (a gene for which the gene-disease relationship is not fully established).^25^ Given the aim of the study was to study clinical management actions from genetic testing results, combining these biologically distinct categories into a single comparator group is methodologically appropriate. Variants included in this study were deposited into the National Institutes of Health ClinVar database.

### Survey design

An 11-item structured quantitative questionnaire, formatted as a clinical report form, was designed using Qualtrics survey software (Qualtrics, Provo, UT) to collect clinical data and management decisions made by cardiologists after conducting a chart review for patients who received either a positive or a non-positive genetic testing result. Survey items were developed by A.M., Y.T., B.B., and E.D.E. (available in Supplement). Participating cardiologists tested the questions and survey platform and then offered suggestions to ensure the clinical appropriateness and clarity of the items and the timely completion of the survey. The survey remained open from September 10, 2024 to December 4, 2024.

For each randomly assigned case, cardiologists were asked “What medical management change(s) did you recommend based on the genetic test result?” (question 10). We grouped responses to question 10 into five broader categories related to the patient’s own care: changes to recommended cardiac procedure, cardiac evaluation, medications, lifestyle, and specialist referral (Supplemental Table S2). Affirmative responses (i.e. responding “Yes”) to at least one of the related questions were counted as a management recommendation for that category.

Management recommendations related to the patient’s family were related to cascade genetic testing or cardiac evaluation. Clinical management recommendations are broadly reported for the overall cohort, while specific management change recommendations are provided for positive vs non-positive cohorts, and for each positive gene.

### Statistical methods

Only completed responses for unique patients were included. Free-text responses were analyzed qualitatively and reconciled. Aggregate quantitative data were summarized using descriptive statistics. Continuous variables are reported as a mean with standard deviations. Categorical variables are reported as a percentage. Chi-squared or Fisher’s exact testing was performed where appropriate to compare groups. Logistic regression was performed to assess the likelihood of receiving a recommendation for the patient’s own management with a positive result in an ACM gene compared to a positive *TTN* result while controlling for patient age, sex, and race and/or ethnicity. Statistical significance was defined as a two-sided *P* value of < 0.05. *P* values were adjusted for multiple comparisons using Bonferroni correction where appropriate.

## RESULTS

### Respondent characteristics

Participating cardiologists who completed the survey practiced in tertiary academic centers. All were board-certified cardiologists, with three also having training in advanced heart failure and transplant and two having electrophysiology training. All respondents reported feeling extremely or somewhat comfortable with identifying patients and interpreting results for CM/ARRH genetic testing. For most genetic testing in their practice, they reported interpreting results in collaboration with other members of the clinical team, which included genetic counselors, nurse practitioners, and other cardiologists (n=4) or by themselves (n=3).

### Cohort description

A final cohort of 249 patients receiving care at 7 different institutions was obtained (Figure 1). The resulting group achieved a sample with 80% power to identify differences in the proportion of patients with positive and non-positive results who received management change recommendations.

Most patients receiving genetic testing were male (n=153, 61.4%), clinician-reported White (n=151, 60.6%), and referred by the treating cardiologist for genetic testing by heart failure (n=104) or electrophysiology (n=63) clinics. Cardiomyopathy was the most common referral reason for genetic testing (n=84, 34%) (Supplement Figure S1). The mean age at genetic testing was 51.9 years (SD=15.7) (Supplemental Figure S2).

Patients with positive results in genes of interest were randomly selected to be in the positive group (n=138) and non-positive patients were randomly selected to be in the non-positive group (n=111) (Figure 2). On average, positive patients were younger than non-positives at testing (50.1 vs 54.6 years, *P* = 0.02). Otherwise, the positive and non-positive cohorts were demographically similar (Table 1).

**Figure 2.**
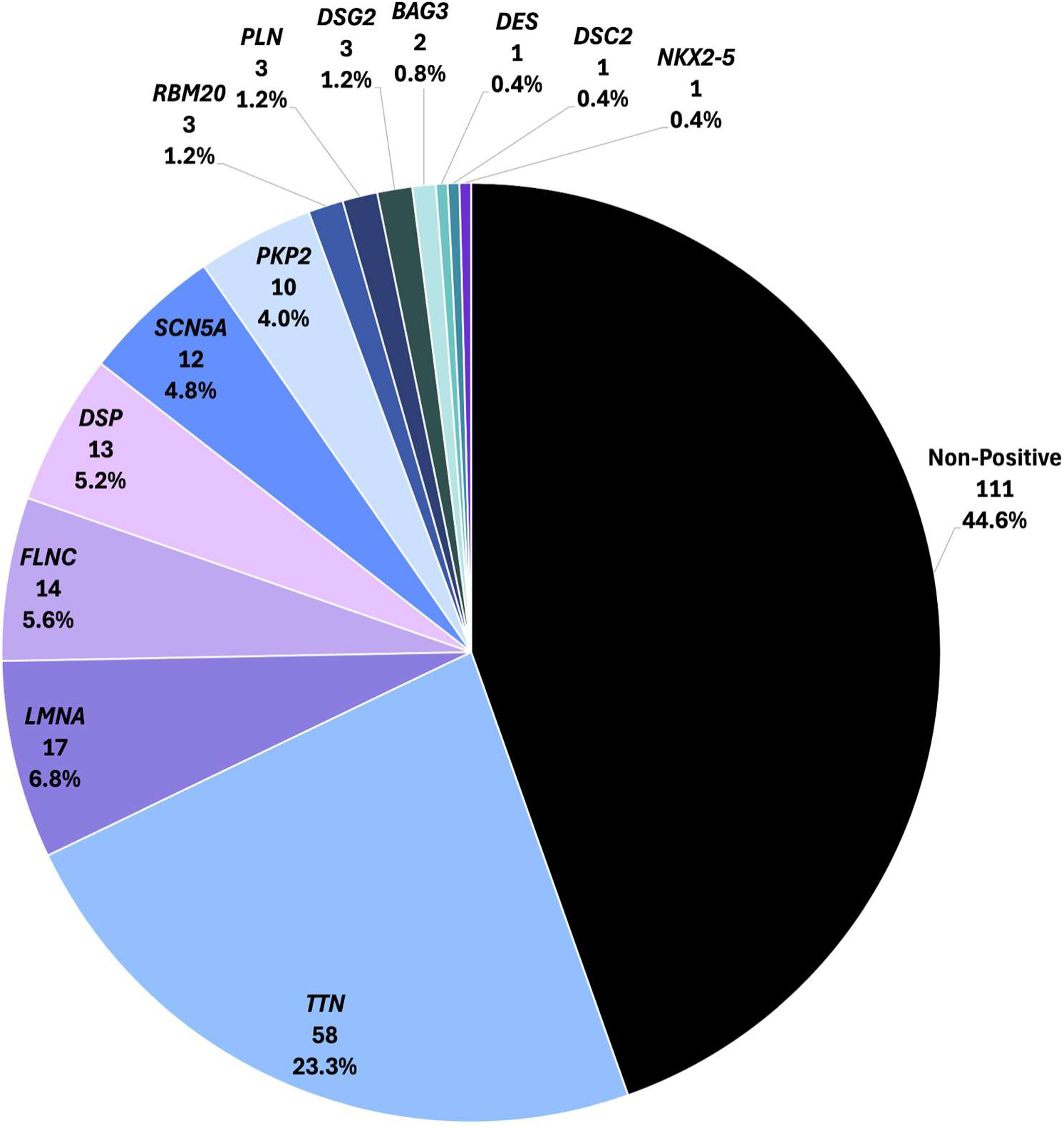
Genetic testing results in the patient cohort. A total of 249 patients were included in the study with positive results in specific arrhythmogenic cardiomyopathy genes of interest (n=138) and non-positive results (n=111). Cardiologists completed case report forms on clinical data and management decisions following the genetic testing result.

**Table 1.**
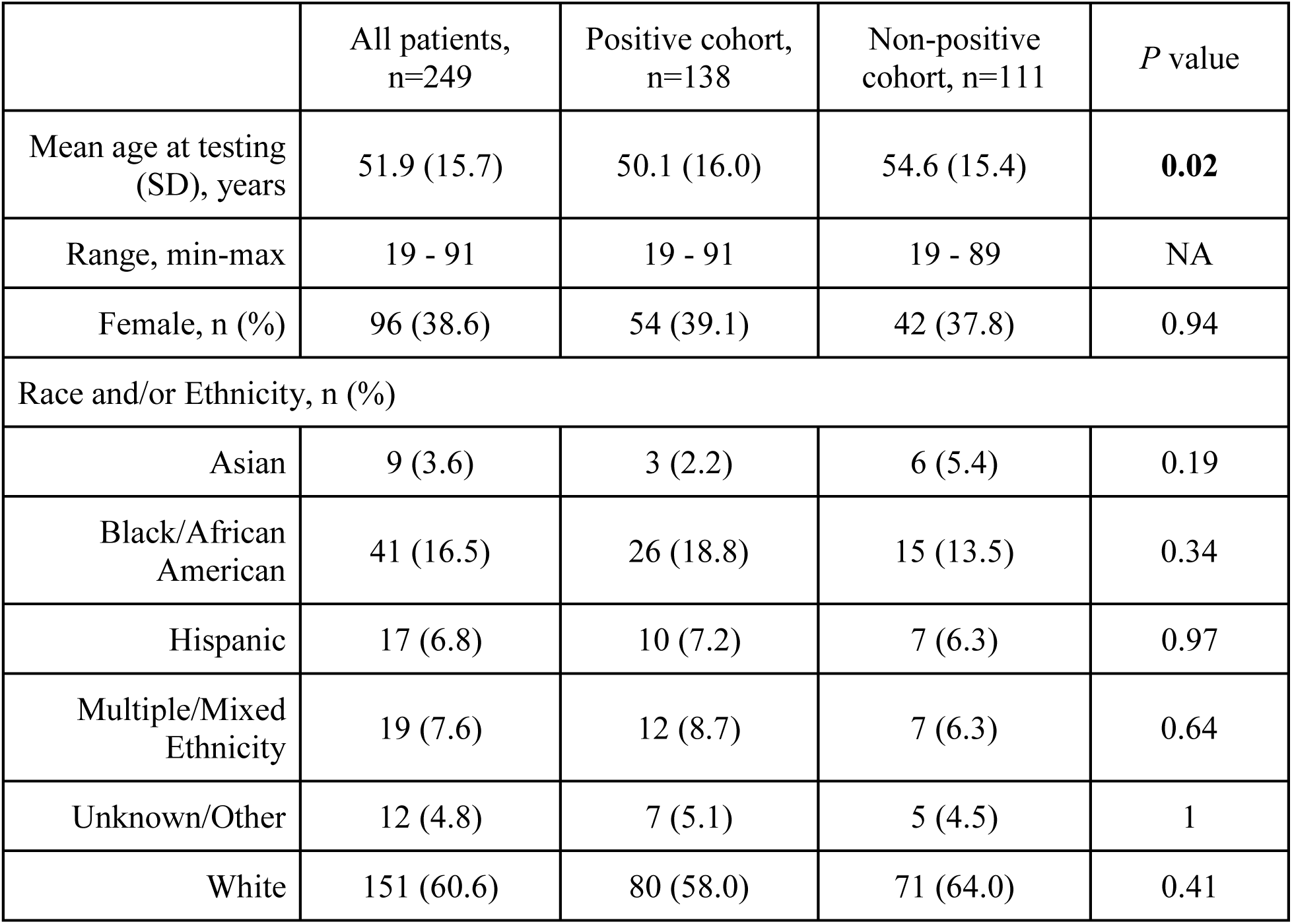
Demographics of patient cohort. Abbreviation: SD = standard deviation.

### Clinical management recommendations

Based on review of their assigned patients’ medical records, participating cardiologists reported that positive and non-positive genetic testing results led to a change in clinical management for 136/249 (54.6%) patients (Figure 3). Of the 136 patients, 75 (55.1%) had management change recommendations made for their own care, consisting of cardiac procedures (n=50), medication (n=40), cardiac evaluations (n=29), lifestyle (n=26), and specialist referral (n=23). Recommendations for family members included cardiac evaluation (n=103) and cascade genetic testing (n=88).

**Figure 3.**
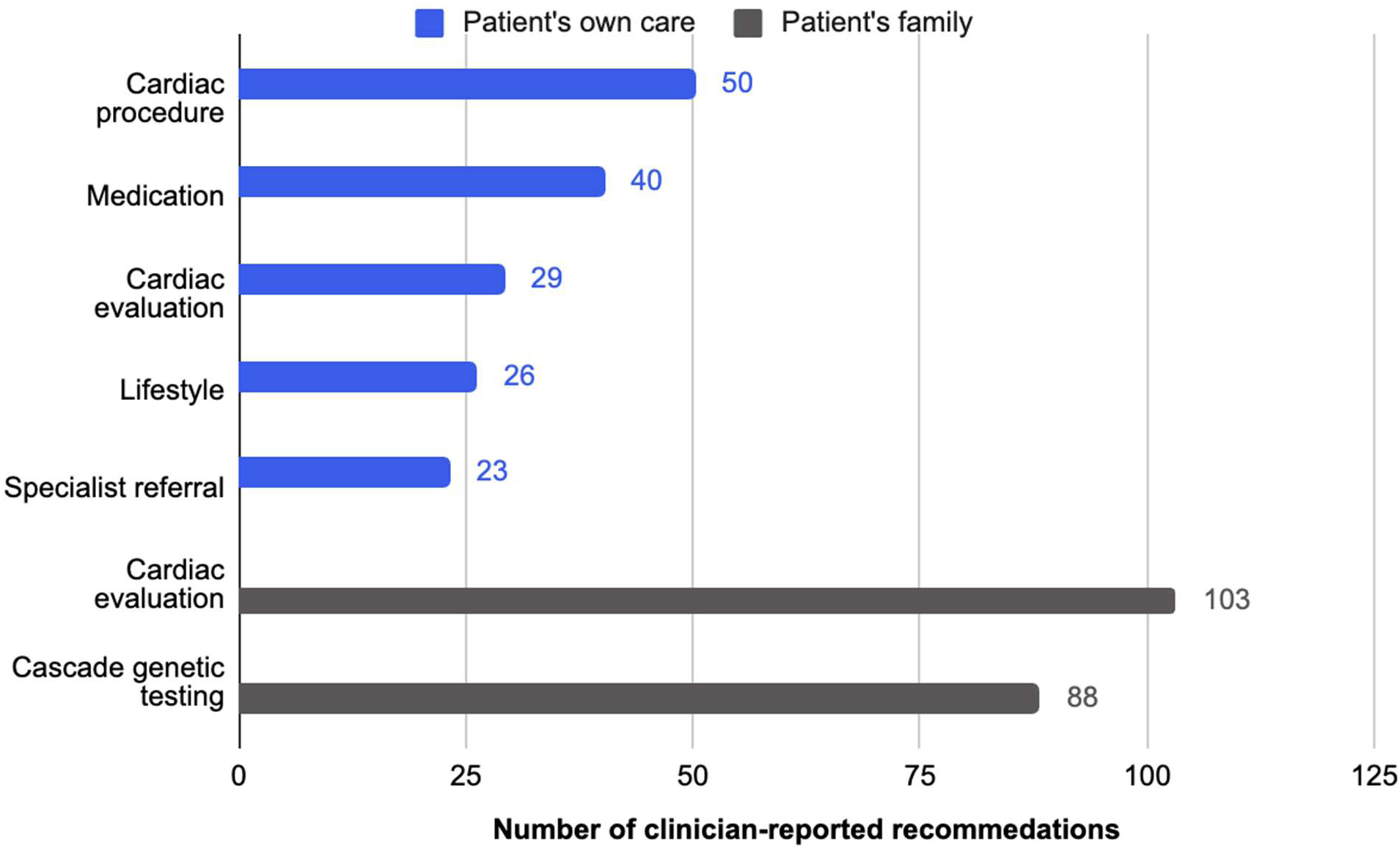
Clinician-reported recommended management changes. Overall management changes recommended in this cohort related to the patient’s own care and their at-risk relatives. Broader categories are grouped based on responses to specific questions from the case report form. Categories and specific medical management changes are listed in Supplemental Table S2.

No changes in clinical management based on genetic testing results were reported for 113 (45.4%) patients. Of these, 71 (62.8%) received no management change recommendations because the genetic testing result was not informative to clinical management. In 40 (35.4%) cases, the patient’s current management plan was already consistent with recommendations based on the genetic testing result. In 2 (1.8%) patients, the patient was deceased or lost to care.

### Comparison of management change recommendations among positive and non-positive genetic testing results

Genetic testing results were associated with management change recommendations for patients with both positive and non-positive results. Overall, 109/138 (79.0%) of positive and 27/111 (24.3%) of those with non-positive results had some type of management change recommended (*P* < 0.0001). Among the 75 patients for whom management change recommendations were made to their own care, a significantly higher proportion of positives received recommendations when compared to the non-positive group (66/138, 47.8% vs 9/111, 8.1%; *P* < 0.00001).

### Clinical management change recommendations for the patient’s own care

#### Positive ACM genes

At least one management change recommendation for the patient’s own care was made for each ACM gene in the positive cohort (Figure 4). Recommendations included diagnostic test/procedure (n=33), referral to a specialist (n=20), and device implantations (n=21) (Figure 5A). Of those with positive ACM genes and recommendations for device implantation, recommendations for implantable cardiac defibrillator (ICD) placement were made for patients with *LMNA* (4/17 carriers), *FLNC* (2/14 carriers), *DSP* (2/13 carriers), *SCN5A* (2/12 carriers), *PLN* (1/3 carriers), and *DES* (1/1 carrier), and a biventricular pacemaker in a patient with *LMNA* (1/17 carriers). Recommendations for extra-cardiac disease monitoring were made for patients with *LMNA* (3/17 carriers), *FLNC* (3/14 carriers), *DSP* (1/13 carriers), *SCN5A* (1/12 carriers), and *DES* (1/1 carrier).

**Figure 4.**
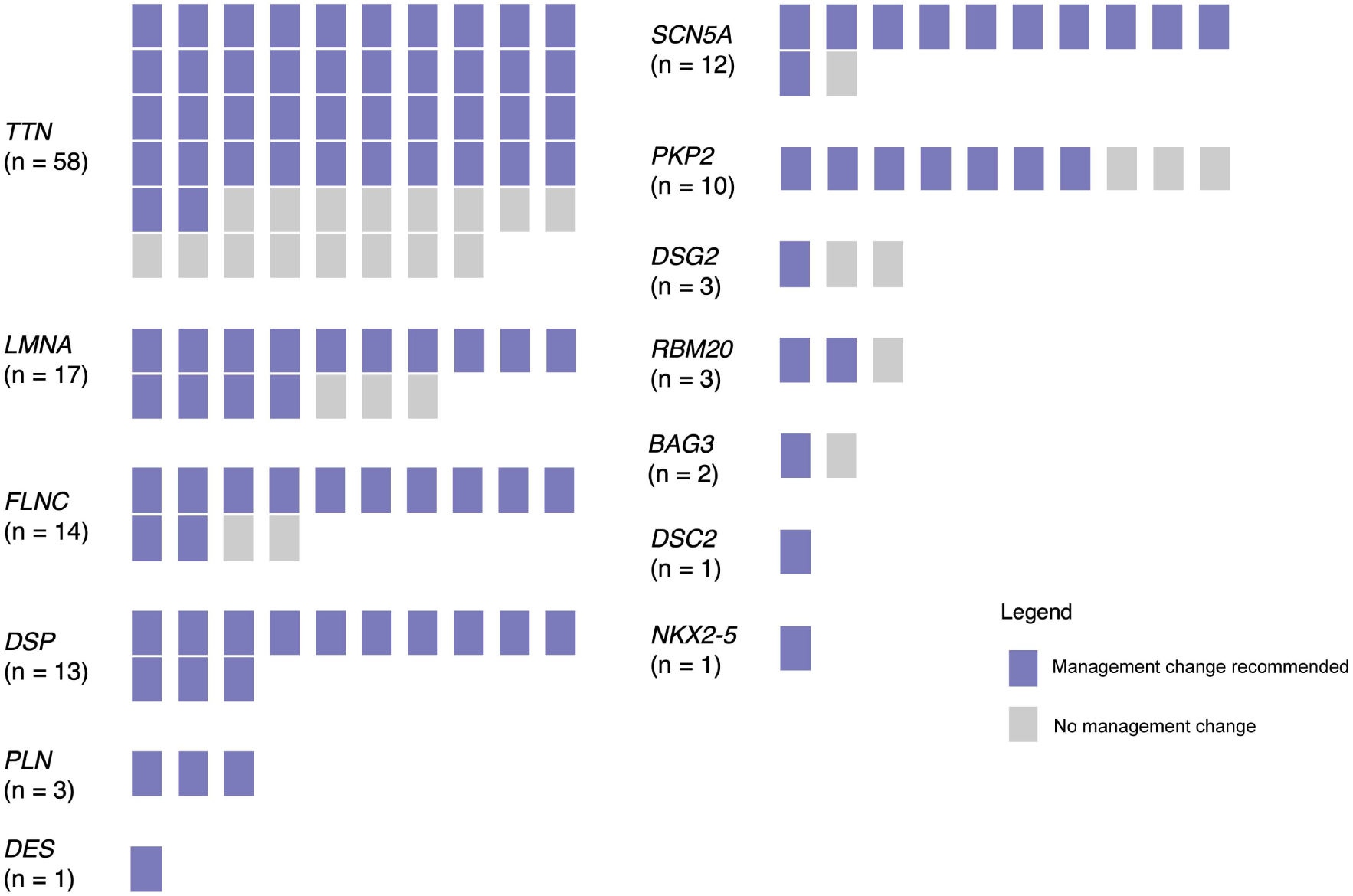
Clinical management changes recommended for patients with positive test results. Cardiologists completed a case report form on genetics-informed clinical management change recommendations. Each rectangle represents a single patient for whom a management change was recommended (purple) or not recommended (grey).

**Figure 5.**
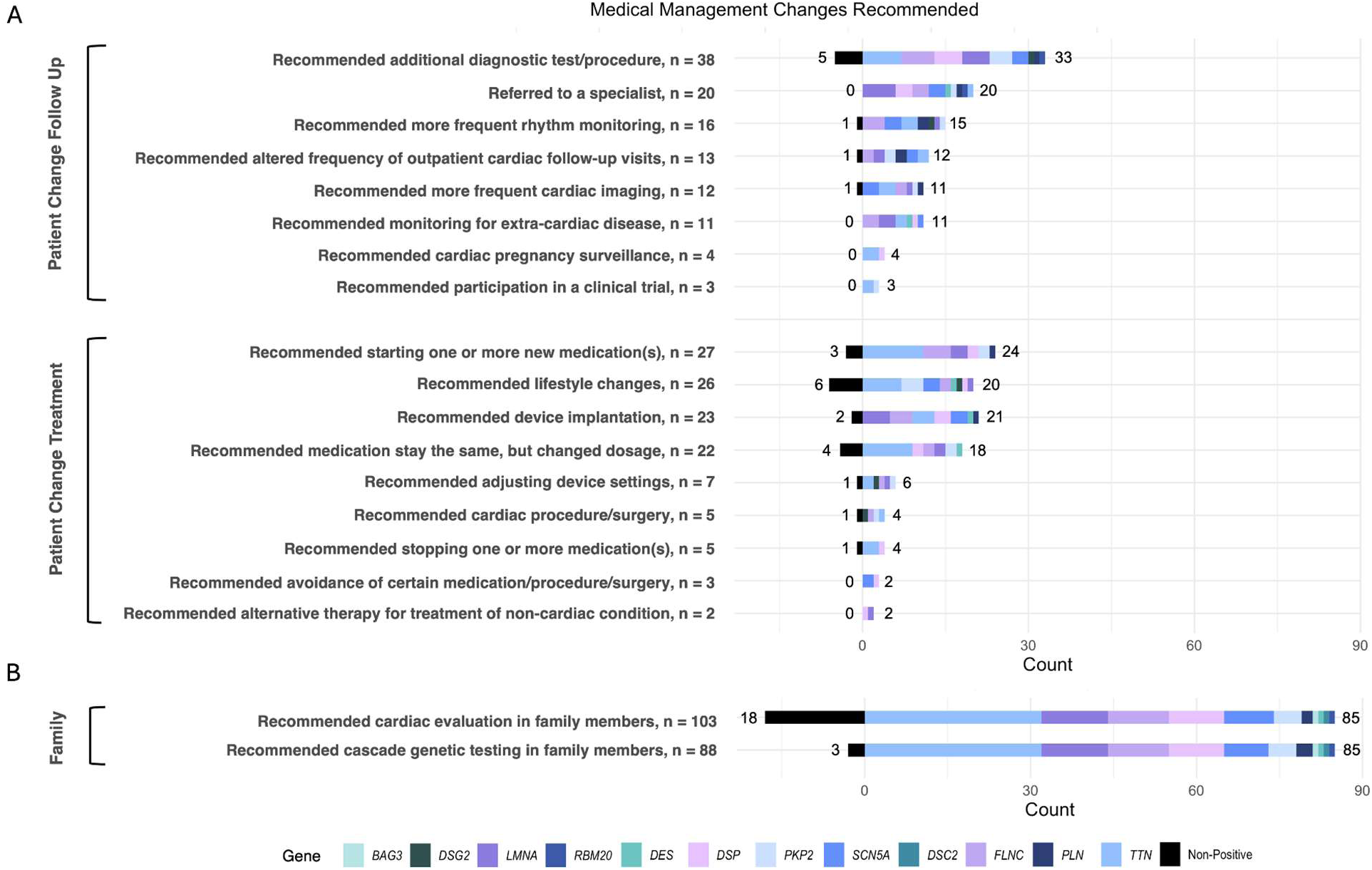
Clinical management changes reported. Cardiologist-reported recommendations following the receipt of genetic testing results for **A**) the patients’ own care and **B**) the patient’s family. Each bar shows the total count of recommendations made. The same patient may be represented in more than one bar since multiple recommendations may be made for a single patient (e.g. referral to a specialist and recommended cardiac evaluation in family members).

#### Positive TTN results

Overall, 20/58 (34.5%) of patients with a positive *TTN* result received management recommendations for the patient’s own care (Figure 4), consisting of starting one or more new medications (n=11), changing medication dosage (n=9), lifestyle changes (n=7), and additional diagnostic procedures (n=1) (Figure 4A). An ICD was recommended in 3/58 *TTN* carriers. Despite patients with a positive *TTN* result accounting for 42.0% (58/138) of the positive cohort, patients with positive results in ACM genes had 263% higher odds of receiving a change in management recommendations compared to those with a positive *TTN* result (OR = 3.63, CI: 1.40 - 9.84, *P* = 0.009).

#### Non-positive results

Patients with non-positive results also received medical management recommendation changes based on their genetic test result. These changes consisted of lifestyle modification (n=6), additional diagnostic tests/procedures (n=5), changing medication dosage (n=4), starting a new medication (n=3), LVAD or ICD implantation (n=2), and recommending a cardiac procedure/surgery, stopping a medication, adjusting device settings, more frequent rhythm monitoring, more frequent cardiac imaging, more frequent outpatient cardiac follow up visits (n=1 each, Figure 5A).

### Clinical management change recommendations for the patient’s family

Cardiac evaluation was recommended for family members of 85/138 (61.6%) of patients with positive and 18/111 (16.2%) of patients with non-positive results (Figure 5B). Cascade genetic testing was recommended for family members of 85/138 (61.6%) of the positive cohort and 3/111 (2.7%) of the non-positive cohort (Figure 5B).

## DISCUSSION

### Result implications

For this retrospective study, participating cardiologists reported using CM/ARRH genetic testing results to inform medical decision making beyond the patient’s genetic diagnosis and family recurrence risk. Of those with recommended management changes, 75/136 (55.1%) of patients received at least one recommendation for their own care. Exploratory studies of potential^1,13^ and probable^14–17,19,20,26^ clinical management changes after CM or ARRH genetic testing have mainly focused on primary diagnosis. To our knowledge, this is the first study to directly query clinicians as to how genetic information specifically altered clinical management. The clinical cohort in this study all received the same genetic test, and clinical recollection was obtained through a standardized case reporting form to document medical decision making.

#### Recommendations for patients with positive results in ACM genes

The survey was designed to capture the medical management change recommended based on the genetic testing result (question 10). While we could not assess the other factors in making a determination for a recommendation (medical history, symptoms, risk monitoring or family history), the recommendations made by participating cardiologists for patients in our cohort with positive ACM genes were consistent with clinical practice guidelines.^27^ These included additional diagnostic test/procedures for 33 patients, ICDs for 15 patients with positive results in *LMNA*, *SCN5A*, *FLNC*, *DSP*, *DES*, and *PLN*, starting a new medication for 24 patients, cardiac procedures for 4 patients, and lifestyle modifications for 20 patients (Figure 5A). Additionally, certain ACM genes may have extra-cardiac presentation (e.g. skeletal myopathy). In our positive ACM cohort, 11 recommendations for extra-cardiac disease evaluation were made for patients with positive results in *LMNA, FLNC, DES*, *DSP*, and *SCN5A*.

#### Recommendations for patients with positive TTN results

*TTN* is the most common genetic cause of cardiomyopathy, and it is also the most frequently identified positive gene in patients referred for genetic testing.^1^ As expected and consistent with guidelines, positive results in ACM genes, rather than *TTN,* were more likely to receive a recommended change.^6^ Nonetheless, 34.5% of those with a positive *TTN* result also received a management change recommendation for the patient’s own care. Individuals with cardiac conduction disease or atrial fibrillation may progress to CM when a disease-causing variant in *TTN* is present,^5,28^ warranting closer monitoring and potentially earlier treatment with guideline-directed medical therapy for heart failure. The results from our study, alongside larger cohort studies of *TTN* carriers^29^ may help inform future studies and consensus regarding management recommendations for patients with positive results in this gene.

#### Recommendations for patients with non-positive results

Interestingly, we uncovered that non-positive results could also result in clinical management for the patient’s own care (Figure 5A). In 5 patients, additional diagnostic tests or procedures were recommended following genetic testing, suggesting genetic testing informed the workup for a “diagnosis of exclusion,” where clinicians perform a process of elimination by excluding possible conditions. For example, a negative genetic test for *DSP* or other genes associated with cardiac inflammation, may direct cardiologists to consider inflammatory cardiomyopathies, which are complex due to their variable presentation.^30^ Just as a positive result may help to limit unnecessary use of other diagnostic testing, such as a positron emission tomography scan or an endomyocardial biopsy, a non-positive result may help to support further investigation for the underlying cause.

### Clinical management change recommendations for the patient’s family

The proportion of patients who received recommendations for cardiac evaluations in their family members (61.6% and 16.2% of the positive and non-positive, respectively) and cascade genetic testing for family members (61.6% and 2.7% of the positive and non-positive, respectively) were lower than expected based on professional societies recommendations for clinical screening^4,6^ and cascade genetic testing for at-risk family members.^4^ For our cohort, it is possible that no living relatives were available, or relatives may already have had genetic testing (making a recommendation redundant). Further investigation about how information about genetic testing is shared within family members is warranted and additional efforts to improve family cascade testing may be needed.

### Timing of genetic testing in patient journey

In this cohort, most patients presented with advanced disease or already had a major cardiac event (e.g. survived cardiac arrest). This suggests the timing at which patients received genetic testing is late in their disease journey. Future studies are needed to evaluate whether the timing of genetic testing is associated with differences in management recommendations and health outcomes. Population-based genomic screening programs and expanded use of the Centers for Disease Control and Prevention including CM/ARRH as a tier 1 genomic application may lead to this type of proactive approach in the future.^31^

### Limitations

The major limitation with this study design is its retrospective nature and potential “recall bias.” Future investigation could use a prospective analysis, but a prospective study design may still suffer from overvaluing the genetic testing result, as clinicians may seek to reinforce their rationale for ordering genetic testing. In our study, we relied on a select group of experienced cardiologists in high volume academic medical centers, where clinicians often have access to robust teams of other cardiology specialists and genetics providers. We relied on genetically-experienced cardiologists in an effort to estimate a “best case” scenario, understanding that genetic testing by frontline cardiologists with less expertise is not likely to have this degree of influence on medical recommendations. Given the minority of eligible patients that currently receive genetic testing,^8,9^ conducting this study across a broad range of expertise may not be sufficient to receive meaningful results. The results represent the practice of cardiologists from 7 institutions, and this number is too small to adjust analyses for individual cardiologists or practice sites. As such, management change recommendations do not reflect clinician-level practice variation. Another limitation is that the cardiologists in this study were invited to participate by querying a database of ordering providers from a single commercial laboratory. Because multi-gene panels from commercial and academic genetic testing laboratories are similar, we do not expect that the source of testing influenced the outcomes.^32^

In conclusion, genetically-experienced cardiologists reported using genetic testing results to inform patient care recommendations. While our findings should be interpreted in the context of the selected gene set, this study documents that genetic testing can lead to management change recommendations beyond confirming a suspected diagnosis and guiding at-risk relatives’ care.

## ACKNOWLEDGEMENTS

None.

## DISCLOSURES

A.M. is a former employee of Invitae (now part of Labcorp).

Y.T., B.B., E.D.E. are current or former employees of Labcorp.

D.P.J. has received consulting payments from Alexion, Alnylam, Attralus, Bayer, BridgeBio

Lexeo Therapeutics, Novo Nordisk, and Rocket Pharmaceuticals.

J.W. is a consultant for Bristol Myers Squibb.

E.D.E. is an advisor and stockholder of Exir Bio and ROMTech.

V.V. has received research grants from Tenaya Therapeutics.

E.M.M. is or was a consultant for PepGen, Tenaya, Novartis, co-founder of Kardigan, and founder of Ikaika Therapeutics.

## SUPPLEMENTAL MATERIALS

Supplemental Table S1. Multi-gene panel genetic testing.

Supplemental Table S2. Broader categorization of management recommendations.

Supplemental Figure S1. Reason for referral for genetic testing.

Supplemental Figure S2. Age of symptom onset and genetic testing.

Supplement. Clinical report form

## ABBREVIATIONS

ACM: arrhythmogenic cardiomyopathy
ARRH: arrhythmia
CM: cardiomyopathy
P/LP: pathogenic/likely pathogenic
SD: standard deviation
VUS: variant of uncertain significance

